# Real-time monitoring of the effectiveness of six COVID-19 vaccines in Hungary in 2021 using the screening method

**DOI:** 10.1101/2022.02.18.22271179

**Authors:** Horváth J. Krisztina, Tamás Ferenci, Annamária Ferenczi, Gergő Túri, Gergely Röst, Beatrix Oroszi

## Abstract

Several studies have reported a waning of the effectiveness of COVID-19 vaccines. We report real-life vaccine effectiveness in Hungary, estimated with the screening method, in 2021, i.e., covering the dominance of both the Alpha and the Delta variant, and including the booster roll-out. Hungary is in the unique position to use six different vaccines (including the Sputnik V and Sinopharm vaccines, for which limited evidence was available prior to the present study) in the same, relatively homogeneous population. All vaccines provided high level of protection initially which declined over time. While the picture is different in each age group, the waning of immunity is apparent for all vaccines and especially in the younger age groups and the Sinopharm, Sputnik-V and AstraZeneca vaccines, which performed similarly. This is clearly reversed by booster doses, more prominent for those vaccines, where recipients were more likely to take the booster dose (which were the aforementioned three vaccines). Booster doses were almost exclusively mRNA vaccines. Overall, two vaccines, Pfizer/BioNTech and Moderna tend to produce the best results in all age groups, and even with waning taken into account.

## Introduction

Several studies have reported a waning of vaccine effectiveness (VE) of COVID-19 vaccines, particularly in the period when the Delta variant became dominant [1, 2], however few countries are in the position to monitor brand specific VE for a so wide range of different vaccines as in Hungary. Hungary launched its COVID-19 vaccination programme on week 52 of 2020 and has deployed an extensive vaccine portfolio since then consisting of six different types of vaccines [3] as shown in Figure 1.

**Fig 1.**
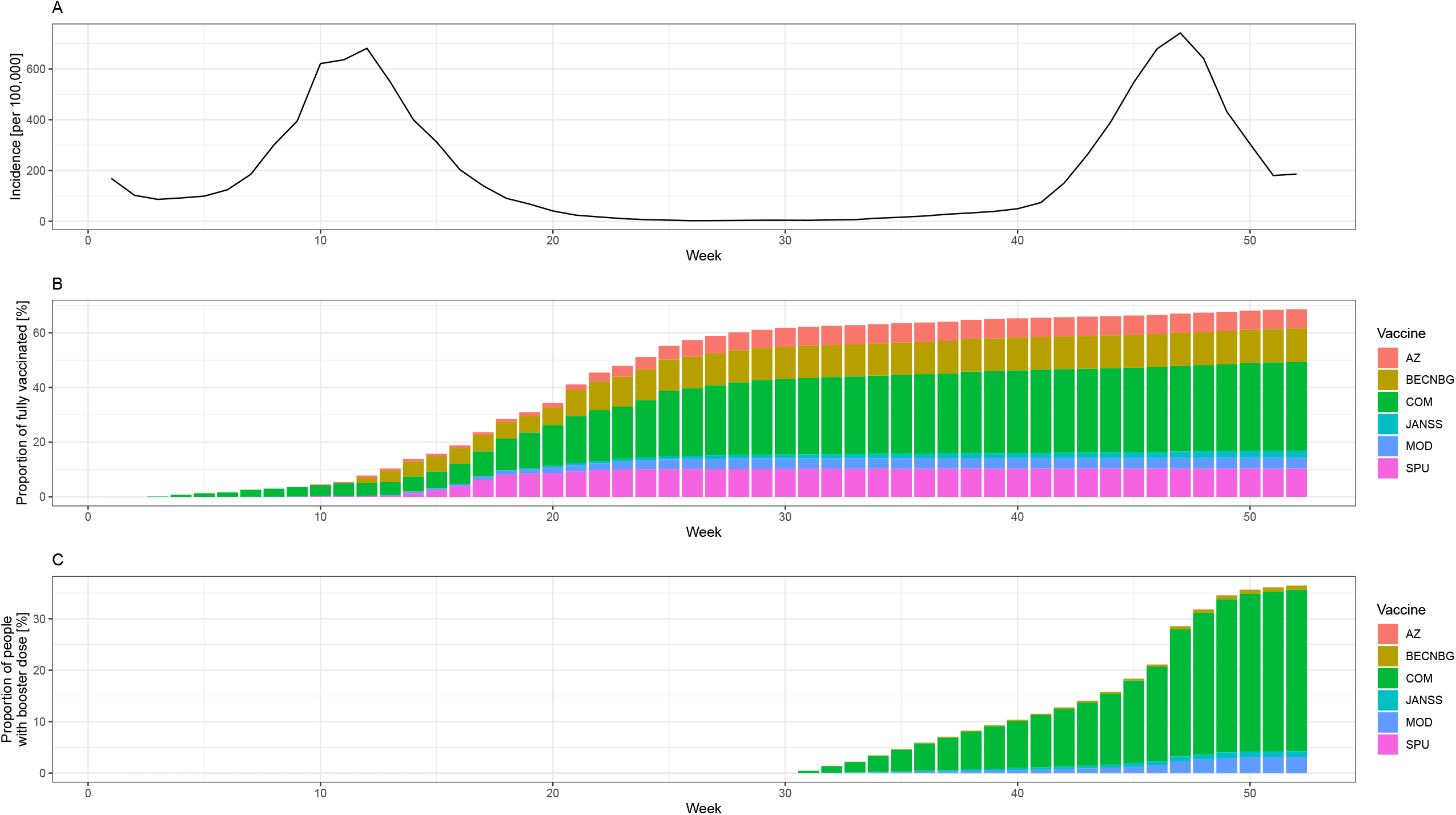
Epidemiological situation in Hungary (weekly number of reported cases per 100,000, panel A), proportion of fully vaccinated people by week and vaccine brand in Hungary (panel B) and proportion of people who received booster dose by week and vaccine brand (panel C) among those aged 12 or more, in 2021. Abbreviations: AZ: ChAdOx-1 (AstraZeneca), BECNBG: BBIBP-CorV (Sinopharm), COM: Comirnaty (Pfizer/BioNTech), JANSS: Janssen, MOD: mRNA-1273 (Moderna), SPU: Gam-COVID-Vac (Sputnik V) vaccine.

The aim of this study is to expand the evidence base about the decreasing protection from COVID-19 vaccines over time, by vaccine brands, covering six vaccines including the Sputnik V and the Sinopharm vaccines (for which limited evidence was available prior to the present study), in the same, relatively homogeneous population, and to report the effectiveness of booster doses in real world settings in a time period when the SARS-CoV-2 Alpha and Delta variant was dominant.

## Materials and Methods

The screening method [4] can be used to estimate VE if information is available on the vaccine uptake of the population and on the proportion of vaccinated among the infected patients. No information is needed however on those who are non-infected – apart from the vaccine coverage – which makes it especially suitable for VE monitoring in low-resource countries, as information on infected patients is much easier to obtain, and is often routinely done anyway.

Data on vaccine uptake, i.e., the weekly number of first, second and third doses administered from each vaccine brand, stratified according to age groups (12-17, 18-24, 25-49, 50-59, 60-69, 70-79, 80+ years of age) were obtained from the European Centre for Disease Control and Prevention (ECDC) [3]. Brand-specific percentage of population vaccinated (PPV) on a given week was defined as the cumulative number of second (every vaccine except Janssen) or first (Janssen) doses administered by the given week from a given brand, divided by the sum of this number and the number of unvaccinated. For the purpose of calculating PPV, unvaccinated was defined as not having received any dose from any vaccine, i.e., the number of unvaccinated was the denominator minus the sum of the cumulative first doses from all vaccine brands. For a given week, the PPV two weeks prior to the week was used as PPV in the analysis.

The Hungarian Notifiable Disease Surveillance System, operated by the National Public Health Center (NPHC) provides data on notified COVID-19 cases in Hungary. NPHC linked each registered laboratory-confirmed SARS-CoV-2 infection to the database of vaccinations, thereby providing both the date and the brand of the first, second and third dose of vaccination – or the lack of thereof – for each infected person. We defined persons as fully vaccinated 14 days after receiving either the second of two recommended doses of a two-dose vaccine or a single dose of the Janssen vaccine, regardless of whether the person received a third booster dose [5]. For the brand-specific analysis, we defined brand based on the brand of the first vaccine (it only occurred in 0.3% of the cases that the brand of the first and second dose differed). Brand-specific percentage of cases vaccinated (PCV) was defined as the number of people fully vaccinated with a given brand divided by the sum of the number of people fully vaccinated with a given brand and the number of unvaccinated, all among those infected in a given week. That is, partially vaccinated people were coherently excluded both from the numerator and the denominator, similar to PPV. For calculating PCV, persons were considered unvaccinated if they did not receive any COVID-19 vaccine or were vaccinated less than 14 days before the day of laboratory confirmation of infection.

From both data sources we extracted information for 2021. The data are still provisional, the latest as of 24^th^ January, 2022 was used. There are recommendations against using the screening method when the vaccine coverage is rapidly changing [5]; we will present results only after week 20, where changes are no longer dramatic (see Figure 1).

VE was modelled with logistic regression, using calendar week, vaccine brand and age group as covariates. Interaction was allowed between all three covariates, which essentially means that separate time trends are allowed for every vaccine brand and age group combination. The regression modelling approach to the screening method was further developed in the present paper, as calendar week was expanded with thin plate regression splines with the aim of realizing an integrated smoothing of time effect that is needed due to the noisy nature of the data (especially at times where the number of infected is low). That is, the whole problem was recast in the framework of Generalized Additive Models [6]. The application of (spline-expanded) calendar week is particularly important to achieve real-time monitoring of the continuously changing VE over time. Further methodological details can be found in Supplementary Text S1.

Full analysis script, including a synthetic dataset which allows the reproduction of the methods presented here and a simulation validation is available at https://github.com/tamas-ferenci/VaccineEffectivenessEstimationScreeningSpline.

Of note, without case-based data, it is not possible to separate the effect of the booster dose from the primary series in a brand-specific manner, as it will be unknown which was the brand of the primary series (from which the number of administered booster doses should be deducted to obtain the number of those without booster). Thus, brand-specific data will be confounded by the administration of booster doses after they are introduced. Data for the isolated effect of primary series (without booster) will be available only age-, but not brand-specifically, as we can assume that the age – in contrast to the brand – is the same in the primary series and the booster.

## Results

The evolution of brand- and age-specific VE estimates over time is shown in Figure 2. The vertical black line indicates Week 33, when the effect of booster doses may first appear, after that the results pertain not only to the primary vaccine indicated, but to an – unknown – combination of the primary vaccine and the booster. Further results, including the numerical values of VE for week 20, 33 and 52 are given in Supplementary Text S2.

**Fig 2.**
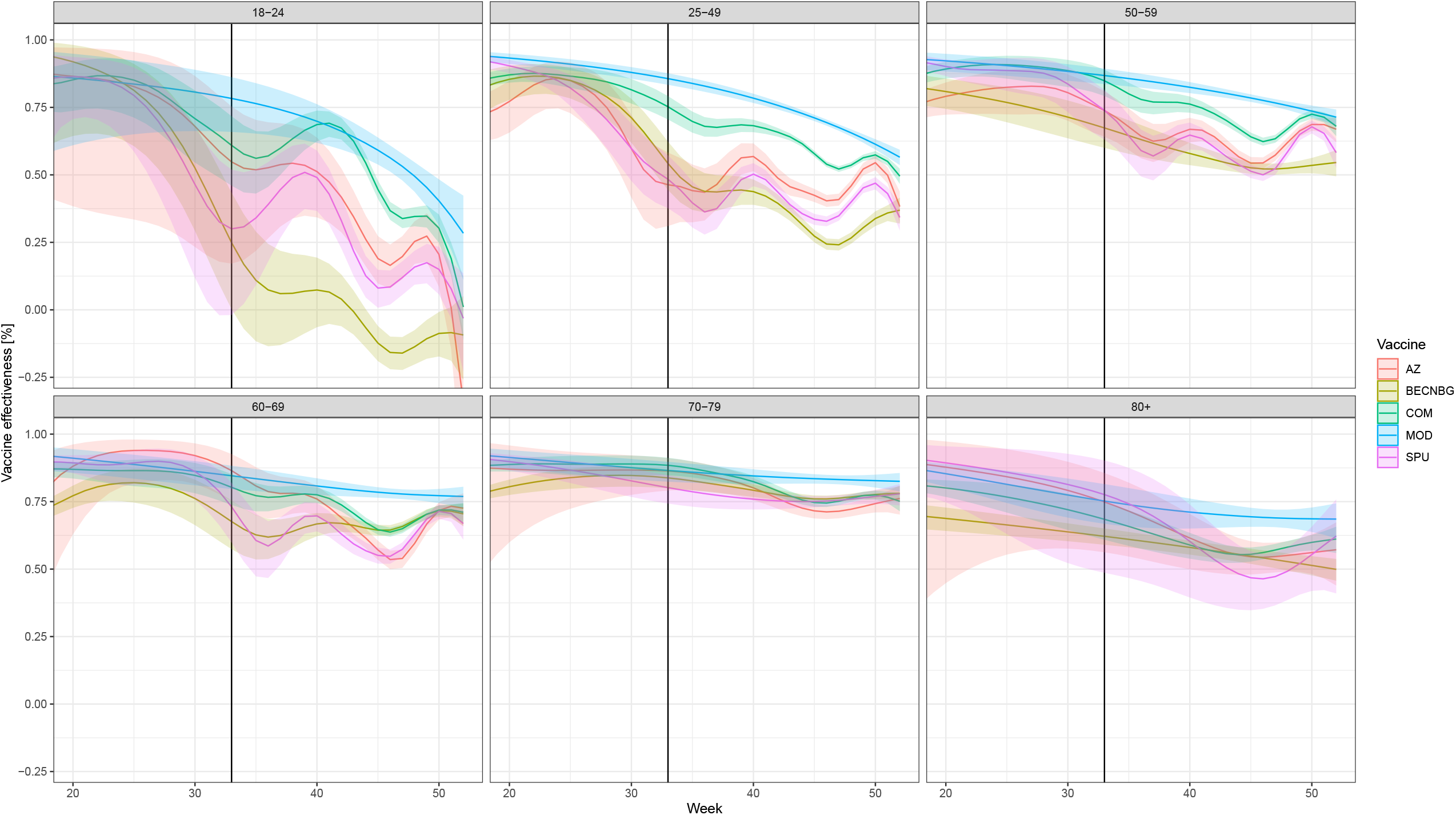
Evolution of VE over time, stratified according to age and vaccine brand in Hungary from week 20 to week 52, 2021. (Due to the low number of people infected who were vaccinated with Janssen vaccine or were below 18, the VEs for these are not presented on Figure, but are given numerically in Supplementary Text S3.) Abbreviations are the same as in Fig 1. Shaded areas indicate 95% confidence intervals. The start of the effect of booster doses from week 33 is shown by a vertical black line; after that the results pertain not only to the primary vaccine indicated, but to an – unknown – combination of the primary vaccine and the booster. Booster doses were almost exclusively mRNA vaccines (Figure 1).

The low number of cases during the summer results in rather wide confidence intervals, but a few overall tendencies are obvious. First, all vaccines provided high level of protection initially which declined over time. While the picture is different in each age group, the waning of immunity is apparent for all vaccines and especially in the younger age groups and the Sinopharm, Sputnik-V and AstraZeneca vaccines, which performed similarly. This is clearly reversed by booster doses, more prominent for those vaccines, where recipients were more likely to take the booster dose (which were the aforementioned three vaccines). Booster doses were almost exclusively mRNA vaccines (Figure 1).

Overall, two vaccines, Pfizer/BioNTech and Moderna tend to produce the best results in all age groups, and even with waning taken into account.

Non brand-specific results for the VE of the primary series only (without booster dose) are shown in Supplementary Text S3.

## Discussion

The continuous, real-time monitoring of VE is crucially important, both for the optimal implementation of public health measures, for informing basic science and for maintaining the trust of the public [7–9].

As for any observational investigation, cohort methods generally provide the best quality evidence, followed by case-control studies. However, if none of them can be implemented, screening method provides a near-real-time, continuous and low resource instrument to survey VE, particularly when denominator data on individual level are not available [4, 5].

The main strength of our study is that it presents evidence on six different vaccines, including the Sputnik and Sinopharm vaccines, for which less information was available prior to this study, in the same, relatively homogeneous population, spanning the entire 2021.

In particular, our study covers the period when the Delta variant was dominant in the EU/EEA, thus it is among the first to provide real world, comparative evidence for the declining vaccine effectiveness of six different vaccines against this variant in the EU/EEA countries. It also gives much needed evidence on the real-life impact of the booster dose.

Importantly, our results are rather similar to those obtained using retrospective cohort design in Hungary in a previous study (HUN-VE) that covered weeks 3-23, 2021, so it may be hypothesized that uncontrolled confounding might not play an important role in our study [10]. The most significant difference from HUN-VE is that our study covers a longer period, including the one when the protective effect of COVID-19 vaccines begins to diminish and includes the phase of booster vaccinations too.

The most important limitation of the present study – in addition to every intrinsic limitation of the screening method itself, especially its particular susceptibility to confounding – is the lack of case-based vaccination data on the receipt of the third those, therefore the inability to control for it.

The second most important limitation is the unavailability of vaccine coverage statistics by more specific risk factors that would allow for more stratification of VE. Selection bias (e.g. enhanced testing for COVID-19 among people with underlying chronic conditions), observer bias (e.g. vaccinated cases might be less likely swabbed) or underreporting also cannot be ruled out in the national surveillance data collection. Moreover, potential confounders such as chronic underlying diseases are not reported, so adjusting for them is not possible. To reduce the impact of uncontrolled confounding, we calculated VE in an age-specific manner, as age is correlated with the presence of comorbidities.

As for any VE study, results might be influenced by the fact that part of the unvaccinated are protected due to prior infection which is not accounted for. This would result in an underestimation of VE, mitigated by the fact that vaccinated also partly gain protection from prior infection.

We had no information on the completeness of linkage between the database on infections and on vaccinations (i.e., how often a missing vaccination data indicates failure to link and not true unvaccinated status).

## Data Availability

The raw data is not publicly available, but the analysis script - with simulated data - is available at https://github.com/tamas-ferenci/VaccineEffectivenessEstimationScreeningSpline.

## Statements and Declarations

### Author contributions

KHJ and TF designed the study and performed the statistical analysis. AF and GT contributed to data collection and drafting the manuscript. GR and BO analysed and interpreted the data. KHJ, TF, AF, GT, GR and BO finalized the manuscript. All authors approved the final version.

### Ethics Approval

The planning, conduct and reporting of the study was in line with the Declaration of Helsinki, as revised in 2013. The study was approved by the Egészségügyi Tudományos Tanács Tudományos és Kutatásetikai Bizottság (Scientific and Research Ethics Committee of the Medical Research Council).

### Consent to Participate

Not applicable.

### Consent to Publish

Not applicable.

## Acknowledgement

The authors acknowledge the Ministry for Innovation and Technology for pro-vision of data of registered laboratory-confirmed SARS-CoV-2 infections including their vaccination status for the Mathematical Modelling and Epidemiology Task Force and thank their continuous support.

