## Supplementary material for "Real-time monitoring of the effectiveness of six COVID-19 vaccines in Hungary in 2021 using the screening method"

**Title**: Real-time monitoring of the effectiveness of six COVID-19 vaccines in Hungary in 2021 using the screening method
**Journal name:** European Journal of Epidemiology
**Authors**: Krisztina Horváth J.[1,2*] , Tamás Ferenci (https://orcid.org/0000-0001-6791-3080) [2,3,4*] , Annamária Ferenczi (https://orcid.org/0000-0002-2597-3867)[1,2] , Gergő Túri (https://orcid.org/0000-0003-3465-8321)[1,2] , Gergely Röst (https://orcid.org/0000-0001-9476-3284)[2,5], Beatrix Oroszi (https://orcid.org/0000-0001-8915-0336)[1,2]
* These authors contributed equally to this work and share first authorship.
**Affiliations**
[1] Epidemiology and Surveillance Centre, Semmelweis University, 25. Üllői Street, 1085 Budapest, Hungary
[2] Mathematical Modelling and Epidemiology Task Force, Hungary
[3] Physiological Controls Research Center, Obuda University, Budapest, Hungary
[4] Department of Statistics, Corvinus University of Budapest, Budapest, Hungary
[5] University of Szeged, Bolyai Institute, Szeged, Hungary
**Corresponding author**: Tamás Ferenci
**Corresponding author information**: Physiological Controls Research Center, Obuda University, Budapest, Hungary.. Address: 1034 Budapest, Bécsi út 96/b. Phone: +36 (1) 666-5553.

### Supplementary Text S1: Derivation of VE in the screening method, regression modeling considerations

Popularized by the landmark publication of C. Paddy Farrington in 1993 [1] and apparently first used in 1980 [2, 3], the screening method (or case-population method) is a pseudo-ecological study. It starts from the usual definition: $VE = 1 - \frac{AR_V}{AR_U}$, where *AR* stands for attack rate (number of infected divided by the size of the population); *V* subscript indicates vaccinated, *U* denotes unvaccinated subjects. By rearranging the terms, it arrives to the following expression:

$$VE = 1 - \frac{AR_V}{AR_U} = 1 - \frac{\frac{I_V}{N_V}}{\frac{I_U}{N_U}} = 1 - \frac{I_V}{I_U} \cdot \frac{N_U}{N_V} = 1 - \frac{I_V}{I - I_V} \cdot \frac{N - N_V}{N_V} = 1 - \frac{I_V/I}{1 - I_V/I} \cdot \frac{1 - N_V/N}{N_V/N}$$

$$= 1 - \frac{PCV}{1 - PCV} \cdot \frac{1 - PPV}{PPV} = 1 - \frac{\frac{PCV}{1 - PCV}}{\frac{PPV}{1 - PPV}},$$

where $I$ and $N$ denote the number of infected and total number of the class indicated by the subscript, respectively.

It is immediately obvious that *1-VE* is an odds ratio. Unsurprisingly therefore, and already noted by Farrington [1], VE can be calculated using a logistic regression, with PCV being the outcome and the logit of PPV used as an offset (the link function is the logit). This approach has two appealing properties, first, a confidence interval is easily obtained, and second, covariates can be included in the regression, which is especially important as it can be used to alleviate the issue of confounding.

The present paper uses the approach with three covariate: calendar week, vaccine brand and age group. The application of calendar week is particularly important, because it is the way to realize a real-time monitoring of the evolution of VE over time. Interaction was allowed between all three covariates, which essentially means that separate time trends are allowed for every vaccine brand and age group combination.

The regression modelling approach to the screening method will also be to some extent further developed in the present paper, as calendar week will be expanded with thin plate regression splines [4] with the aim of realizing an integrated smoothing of time effect that is needed due to the noisy nature of the data (especially at times where the number of infected is low). That is, the whole problem will now be recast in the framework of Generalized Additive Models [5].

Calculations were carried out under the R statistical program package version 4.1.2 [6] using package mgcv version 1.8-38 [5].

Full analysis script, including a synthetic dataset which allows the reproduction of the methods presented here and a simulation validation is available at https://github.com/tamas-ferenci/VaccineEffectivenessEstimationScreeningSpline.

### Supplementary Text S2: Further results of the brand-specific analysis

By week 52 of 2021, the cumulative number of fully vaccinated people was 5,932,375 (68.6%) in Hungary. Breakdown of this figure according to age and vaccine brand is shown on Supplementary Table 1.

| | 12-17 | 18-24 | 25-49 | 50-59 | 60-69 | 70-79 | 80+ |
|---|---|---|---|---|---|---|---|
| **AZ** | 0 | 27451 | 226670 | 176310 | 100722 | 59594 | 19596 |
| **BECNBG** | 0 | 72937 | 328041 | 95937 | 254262 | 249429 | 57914 |
| **COM** | 308946 | 181095 | 1009886 | 361166 | 406863 | 327011 | 214016 |
| **JANSS** | 0 | 24380 | 118297 | 38871 | 20719 | 7391 | 3152 |
| **MOD** | 542 | 18884 | 135999 | 58003 | 58766 | 43218 | 31861 |
| **SPU** | 0 | 54658 | 403973 | 186566 | 180572 | 61043 | 7634 |
| **Denominator** | 589350 | 741094 | 3490565 | 1235206 | 1295058 | 859765 | 438790 |

**Supplementary Table 1. Cumulative number of fully vaccinated people as of week 52 of 2021, according to age (columns) and vaccine brand (rows).**

Numerical vaccine effectiveness estimates for all six vaccines for weeks 20, 33 (first week before booster doses) and 52 are given in Supplementary Table 2, 3 and 4.

| | 12-17 | 18-24 | 25-49 | 50-59 | 60-69 | 70-79 | 80+ |
|---|---|---|---|---|---|---|---|
| **AZ** | NA | 86.5 (38.4-97.1) | 77.2 (65.7-84.9) | 79.1 (72.3-84.2) | 88.1 (62.9-96.2) | 87.1 (60-95.8) | 87.8 (44.8-97.3) |
| **BECNBG** | NA | 91.9 (64.4-98.2) | 85.4 (77.8-90.4) | 80.9 (74.5-85.7) | 77.2 (72.7-80.9) | 80.5 (77.5-83.2) | 68.8 (64.1-72.8) |
| **COM** | NA | 85.2 (73.7-91.7) | 87.1 (84.2-89.4) | 89.2 (85.7-91.9) | 87 (83.6-89.7) | 88.8 (86.2-90.9) | 80.1 (76.7-82.9) |
| **JANSS** | NA | 80.2 (59.5-90.3) | 82.4 (47-94.2) | 85.5 (30.1-97) | 84 (63.7-93) | 87.8 (7.2-98.4) | -2.5 (-1457.3-93.3) |
| **MOD** | NA | 85.9 (61-94.9) | 93.3 (91.1-94.9) | 92.2 (88.3-94.8) | 91.1 (85.7-94.4) | 91.4 (87.4-94.1) | 85.3 (80.6-88.9) |

| | | | | | | | |
|---|---|---|---|---|---|---|---|
| **SPU** | NA | 86.4 (70.7-93.7) | 90.4 (87.6-92.5) | 90.1 (86.6-92.7) | 89.4 (85.5-92.2) | 89.8 (86.5-92.3) | 89.4 (74.8-95.5) |

**Supplementary Table 2. Estimated vaccine effectiveness for different vaccines (rows) and age groups (columns) on Week 20, 2021. 95% confidence interval in parenthesis. NA: vaccine not given in the age group.**

| | **12-17** | **18-24** | **25-49** | **50-59** | **60-69** | **70-79** | **80+** |
|---|---|---|---|---|---|---|---|
| AZ | NA | 54.8 (17.1-75.3) | 46.4 (31-58.3) | 73.9 (64.1-80.9) | 86.5 (75.2-92.6) | 86.4 (79.1-91.1) | 74.9 (56.2-85.6) |
| BECNBG | NA | 24.9 (0.2-43.4) | 54.2 (44.4-62.3) | 67.4 (60-73.4) | 67.6 (57.4-75.4) | 83.8 (80-86.9) | 62.2 (59-65) |
| COM | 90.6 (80-95.6) | 60.9 (46-71.7) | 75.2 (70.5-79.2) | 84.9 (79.2-89) | 80.3 (73.8-85.2) | 88.5 (84.8-91.2) | 68.5 (60.9-74.6) |
| JANSS | NA | 68.3 (53.9-78.3) | 63.3 (47.8-74.2) | 86.5 (76.6-92.2) | 76.2 (63.3-84.5) | 83.5 (62.9-92.7) | 28.3 (-129.7-77.6) |
| MOD | 100 (-Inf-100) | 78.3 (65.9-86.2) | 85.7 (83.4-87.6) | 86.8 (83.7-89.4) | 84.6 (79.7-88.3) | 86.5 (82.4-89.7) | 75.2 (66.8-81.5) |
| SPU | NA | 30 (-1.9-51.9) | 48.4 (37.6-57.2) | 73.9 (63.1-81.5) | 73.1 (59.8-82) | 80.2 (74.3-84.7) | 77.6 (48.6-90.2) |

**Supplementary Table 3. Estimated vaccine effectiveness for different vaccines (rows) and age groups (columns) on Week 33, 2021. 95% confidence interval in parenthesis. NA: vaccine not given in the age group.**

| | **12-17** | **18-24** | **25-49** | **50-59** | **60-69** | **70-79** | **80+** |
|---|---|---|---|---|---|---|---|
| **AZ** | NA | -34.3 (-66.7--8.3) | 38.3 (32.2-43.8) | 66.9 (62.1-71.1) | 72.6 (65.8-78.1) | 76.2 (70.2-81) | 57.2 (43.9-67.3) |
| **BECNBG** | NA | -9.3 (-25.9-5.1) | 36.9 (32-41.6) | 54.6 (49.5-59.2) | 70.5 (66-74.3) | 78.1 (75-80.7) | 49.9 (45.7-53.8) |
| **COM** | 59.6 (52.3-65.8) | 1.1 (-11.4-12.1) | 49.5 (46.6-52.3) | 68 (64.4-71.3) | 71.1 (67.3-74.4) | 75.1 (71.4-78.3) | 61.1 (56-65.7) |
| **JANSS** | NA | 37.1 (25.8-46.7) | 58.9 (52.9-64.1) | 59.7 (51.4-66.6) | 57.2 (48.3-64.6) | 54.7 (36.5-67.6) | -55.8 (-147.3-1.8) |
| **MOD** | 100 (-Inf-100) | 28.4 (11.1-42.3) | 56.6 (53.6-59.3) | 71.4 (68.2-74.2) | 76.9 (72.6-80.5) | 82.5 (78.8-85.6) | 68.5 (61.4-74.4) |
| **SPU** | NA | -3.1 (-23-13.5) | 34.2 (29.4-38.8) | 58.2 (52.6-63.2) | 66.9 (60.9-72) | 77.9 (73.6-81.5) | 62.2 (41-75.8) |

**Supplementary Table 4. Estimated vaccine effectiveness for different vaccines (rows) and age groups (columns) on Week 52, 2021. 95% confidence interval in parenthesis. NA: vaccine not given in the age group. Note that vaccine brand was defined based on the first**

**vaccine, so results presented here pertain to the combined effectiveness of the primary series with the indicated vaccine, and a booster dose (which was received in an unknown proportion, and was almost always an mRNA vaccine).**

#### Supplementary Text S3: Results of the non brand-specific analysis of the VE of the primary series only, without booster dose

Supplementary Figure 1 shows the age- (but not brand-) specific evolution of VE, considering only the primary series (i.e., those who received the third dose are excluded both from the numerator and the denominator).

The figure now shows, as expected, constant waning.
