## Supplementary figures and images for "Real-time monitoring of the effectiveness of six COVID-19 vaccines in Hungary in 2021 using the screening method"

### Supplementary Figure 1

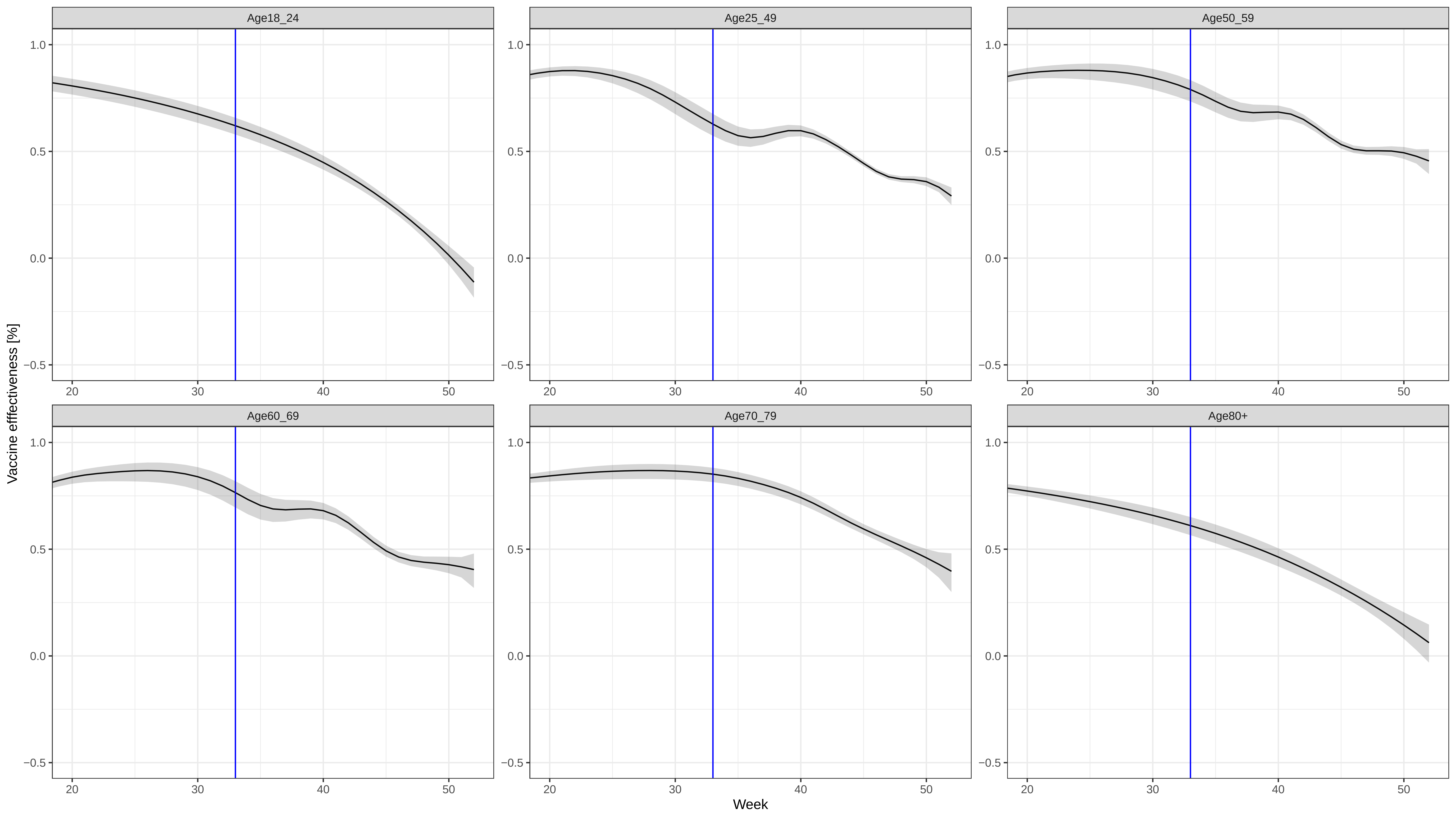
